# Optimal test-assisted quarantine strategies for COVID-19

**DOI:** 10.1101/2020.11.06.20222398

**Authors:** Bo Peng, Wen Zhou, Rowland W. Pettit, Patrick Yu, Peter G. Matos, Alexander L. Greninger, Julie McCashin, Christopher I. Amos

## Abstract

**Objective:** To evaluate the effectiveness of SARS-CoV-2 testing on shortening the duration of quarantines for COVID-19 and to identify the most effective choices of testing schedules.

**Design:** We performed extensive simulations to evaluate the performance of quarantine strategies when one or more SARS-CoV-2 tests were administered during the quarantine. Simulations were based on statistical models for the transmissibility and viral loads of SARS-CoV-2 infections and the sensitivities of available testing methods. Sensitivity analyses were performed to evaluate the impact of perturbations in model assumptions on the outcomes of optimal strategies.

**Results:** We found that SARS-CoV-2 testing can effectively reduce the length of a quarantine without compromising safety. A single RT-PCR test performed before the end of quarantine can reduce quarantine duration to 10 days. Two tests can reduce the duration to 8 days, and three highly sensitive RT-PCR tests can justify a 6-day quarantine. More strategic testing schedules and longer quarantines are needed if tests are administered with less sensitive RT-PCR tests or antigen tests. Shorter quarantines can be utilized for applications that tolerate a residual post-quarantine transmission risk comparable to a 10-day quarantine.

**Conclusions:** Testing could substantially reduce the length of isolation, reducing the physical and mental stress caused by lengthy quarantines. With increasing capacity and lowered costs of SARS-CoV-2 tests, test-assisted quarantines could be safer and more cost-effective than 14-day quarantines and warrant more widespread use.

**RESEARCH IN CONTEXT:** *What is already known on this topic?:*

- Recommendations for quarantining individuals who could have been infected with COVID-19 are based on limited evidence.
- Despite recent theoretical and case studies of test-assisted quarantines, there has been no substantive investigation to quantify the safety and efficacy of, nor an exhaustive search for, optimal test-assisted quarantine strategies.

**What this study adds:**

- Our simulations indicate that the 14-day quarantine approach is overly conservative and can be safely shortened if testing is performed.
- Our recommendations include testing schedules that could be immediately adopted and implemented as government and industry policies.

**Role of the Funding Source:** A major technology company asked that we perform simulations to understand the optimal strategy for managing personnel quarantining before forming cohorts of individuals who would work closely together. The funding entity did not influence the scope or output of the study but requested that we include antigen testing as a component of the quarantining process. Patrick Yu and Peter Matos are employees of Corporate Medical Advisors, and International S.O.S employs Julie McCashin. Other funding sources are research grants and did not influence the investigation.

## Introduction

Until herd immunity to COVID-19 develops through infection or vaccination, quarantine will remain the primary means of disease mitigation. The United States Centers for Diseases Control and Preventions (CDC) currently recommends a 14-day prophylaxis quarantine for anyone who comes into contact with a person who has COVID-19 (1). The 14-day quarantine recommendation is based on clinical data of the observed incubation period (2). Projections indicated that a 14-day quarantine period sequesters 99% of individuals who have been exposed and would ultimately develop COVID-19.

The CDC guideline on a 14-day quarantine has been adopted globally by most organizations and government agencies. Government agencies use quarantine after travel to reduce the introduction of potentially infected individuals into communities.

Industry and organizations, such as professional sports teams and movie productions, have used quarantine to produce a COVID-19-free cohort to resume operations, as mandated by most governments.

Currently, jurisdictions are starting to loosen quarantine requirements due to perceived lower risk of infection, negative psychological effects caused by lengthy quarantines (3), the costs of managing quarantines, loss of productivity, and the need to ensure continuity of operations of essential functions of critical infrastructures. For example, the state of New York announced on October 31, 2020, new COVID-19 travel guidelines that allow travelers to exit quarantine after they test negative four days post-arrival if they also test negative three days prior to departure. As of September 11, 2020, the CDC’s guidance allows critical infrastructure workers to return to work without undergoing quarantine following workplace <u>exposure</u>. The European CDC (ECDC) published <u>revised quarantine requirements</u> allowing quarantine to be discontinued if the result from a SARS-CoV-2 RT-PCR test is negative on day ten after the last exposure with the warning of “residual risk” for ending quarantine early. On December 2, 2020, CDC released a <u>guideline</u> on options to reduce quarantine for contacts of persons with SARS-CoV-2 infection using symptom monitoring and diagnostic testing. The policy allows shortening quarantine to 10 days without testing, or seven days with a SARS-CoV-2 test before release under the conditions of continued monitoring through day 14. The residual post-quarantine transmission risk (PQTR) for a 7-day quarantine with RT-PCR testing was estimated to be 4.0% with a range between 2.3% and 8.6%, based on currently unpublished results (4, 5)

The recommendations from the European and U.S. CDCs reflect both the benefits and uncertainties related to test-assisted quarantine. SARS-CoV-2 RT-PCR was initially developed for diagnostic purposes and later adopted for widespread screening of asymptomatic populations. As RT-PCR can detect very low viral loads, testing individuals before and during quarantine could identify individuals with sub-clinical disease before being infectious, improving quarantine efficacy. Faster and less sensitive antigen tests could potentially also be used during quarantine. These methods were not recommended to the public at the early stages of the pandemic due to insufficient testing capacity and a lack of rigorous assessment of whether testing can sufficiently compensate for the elevated risks related to shortened quarantines. A few recent but non-peer-reviewed studies indicate the benefits of COVID-19 tests during quarantine (4–6).

Nevertheless, test-assisted quarantine has been performed, especially for industries where the benefits of shortened quarantine outweigh the costs of SARS-CoV-2 testing. For example, for movie productions, we have previously found a quarantine strategy with a 3-day pre-travel quarantine, and a 5-day post-travel quarantine with RT-PCR testing provides minimal risk for employees who are forced to work closely together. For individuals who arrived late or had to travel and rejoin the production, more rapid clearance protocols included daily or twice daily testing for five days were recommended. The proposed strategy for movie production was accepted by the cities of Chicago and Atlanta but denied by other governmental agencies. The recommendation and reservations of ECDC’s guidelines and the mixed responses from governmental agencies support the need for a thorough investigation of the risks associated with test-assisted quarantine strategies and the potential to use tests with lower sensitivity, such as antigen testing, for managing cohorts of individuals undergoing quarantines.

Using an individual-based forward-time simulation method (7), we investigated the potential use of SARS-CoV-2 tests to implement a shorter and more effective quarantine strategy. Based on high-sensitivity RT-PCR results, we identified situations where a substantially shorter quarantine period is adequate to mitigate risk for assembling cohorts of protected individuals who will be closely interacting. We recommend guidelines for test-assisted quarantines using widely utilized testing methods and tests with well-characterized analytic and clinical sensitivities, e.g., RT-PCR and rapid antigen tests. We also assessed factors that could affect the performance of test-assisted quarantines via simulation. Here analytical sensitivity refers to the smallest amount of analyte (SARS-CoV-2 virus) in which a SARS-CoV-2 test will be positive for 95% of the time, i.e., the limit of detection (LOD), and clinical sensitivity refers to the proportion of positive index tests in patients who have the disease in question. We noted that clinical sensitivity could vary due to human, specimen, and collection factors, even for the same test kits.

## METHODS

### Simulation method

We used a COVID-19 outbreak simulator to simulate the quarantine of a large number of individuals of varying infectivity patterns (7). While the simulator was designed to simulate the spread of the SARS-CoV-2 virus in a population, we modified it to simulate the quarantine of a large number of individuals, compare the safety of various quarantine strategies, and identify approaches with the shortest quarantine times and least number of tests. We ignored uninfected individuals in our simulations because they do not help evaluate the performance of quarantine strategies. An advantage of this approach was that it did not require any specific analytical form to evaluate viral spread and how interventions could affect transmission.

As depicted in Figure 1, simulated individuals were infected with the SARS-CoV-2 virus either right before entering quarantine or earlier but were asymptomatic when they entered quarantine. Using stochastic models for viral load and transmissibility described below and summarized in Table 1, we simulated the viral load and transmissibility throughout the course of infection. Further, we simulated the times when individuals become infectious, show symptoms if symptomatic, infect others within or outside the quarantine period, and recover (i.e., are no longer contagious but could have a detectable viral load). SARS-CoV-2 tests of specified sensitivity, specificity, and turnaround time were applied, and individuals who tested positive or showed symptoms during the quarantine were removed from the simulation. We estimated post quarantine transmission risk (PQTR) from the proportion of simulations in which the individual completed quarantine and caused one or more infection events after being released.

**Table 1:** Summary of assumptions for the progression and infectivity of SARS-CoV-2 infection.

| Model | Parameter | Value |
| --- | --- | --- |
| **Symptomatic** | R0 | Normal(2.10, 0.36) |
|  | Incubation period | LogNormal(1.62, 0.42) |
|  | Duration of Infection after onset of symptoms | 2 + LogNormal(0.87, 1.03) |
|  | Infectivity Starts | 1/5 of incubation period |
|  | Infectivity Peaks | 2/3 of incubation period |
| **Asymptomatic** | R0 | Normal(0.42, 0.07) |
|  | Duration of infection | 3 + LogNormal(0.97, 0.50) |
|  | Infectivity starts | 1/10 of duration of infection |
|  | Infectivity peaks | 4/10 of duration of infection |

Models used to model the progression and infectivity of SARS-CoV-2 infection for symptomatic and asymptomatic cases. The communicable period of symptomatic cases includes an incubation period modeled by a log normal distribution, and a period of infection after onset of symptoms, which is modeled by a shifted log normal distribution. The communicable period of asymptomatic carriers follows a shifted log normal distribution. Transmissibility probabilities are modeled by piecewise linear functions with a period with no infectivity, followed by a period with increasing infectivity, and then a period of decreasing infectivity.

**Figure 1:**
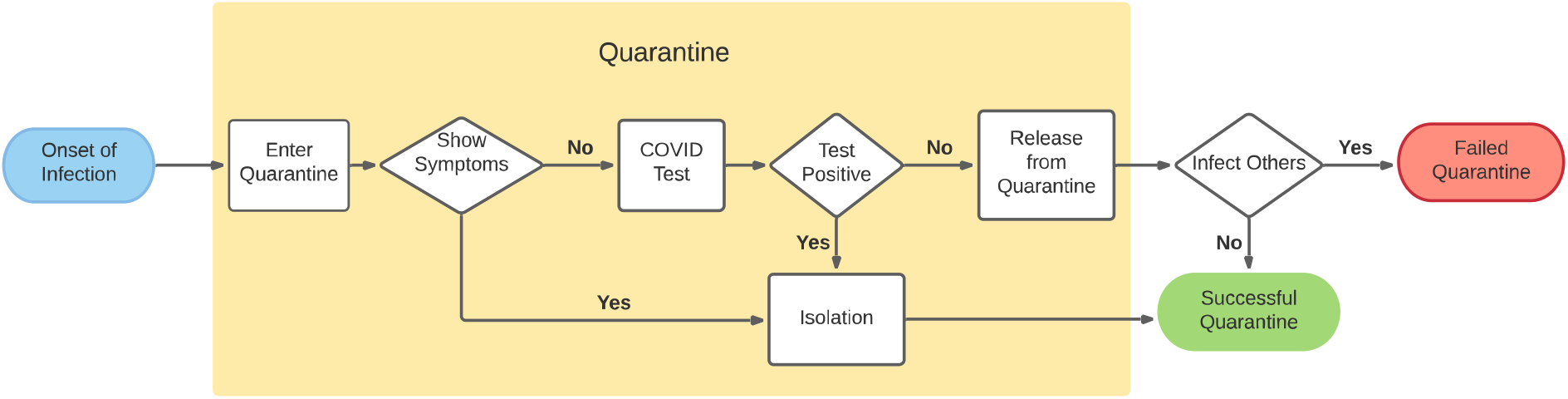
Diagram of a quarantine process with one test performed before the end of quarantine. In our simulations zero, one, or more tests could be performed, and the carrier could show symptoms any time before the end of quarantine and be removed.

Differing strategies may be needed for specific quarantine applications. We simulated scenarios in which individuals are quarantined after either simultaneous infection or exposure to SARS-CoV-2 or after infection at mixed intervals but without symptoms. These scenarios correspond to two major quarantine applications: 1) quarantine of a group of people after simultaneous exposure to a common source of potential infection, e.g., shared close contact with an infected individual or participation in an event with known SARS-CoV-2 positive attendees and 2) pre-quarantine of individuals with an unknown stage of infection before their assembly event, e.g., sporting events, business meetings.

### Model of transmissibility

A transmissibility model determines how many and when infected individuals transmit SARS-CoV-2 to others. We modeled the transmissibility of infected individuals using a piecewise function that starts with a period of non-infectivity, continues with a period of increasing infectivity and concludes with a period of declining infectivity. The overall transmissibility (i.e., the probability of transmissibility integrated over the course of infection) was equal to the reproduction number (R_e_) of the infected individual (Figure 2A).

**Figure 2:**
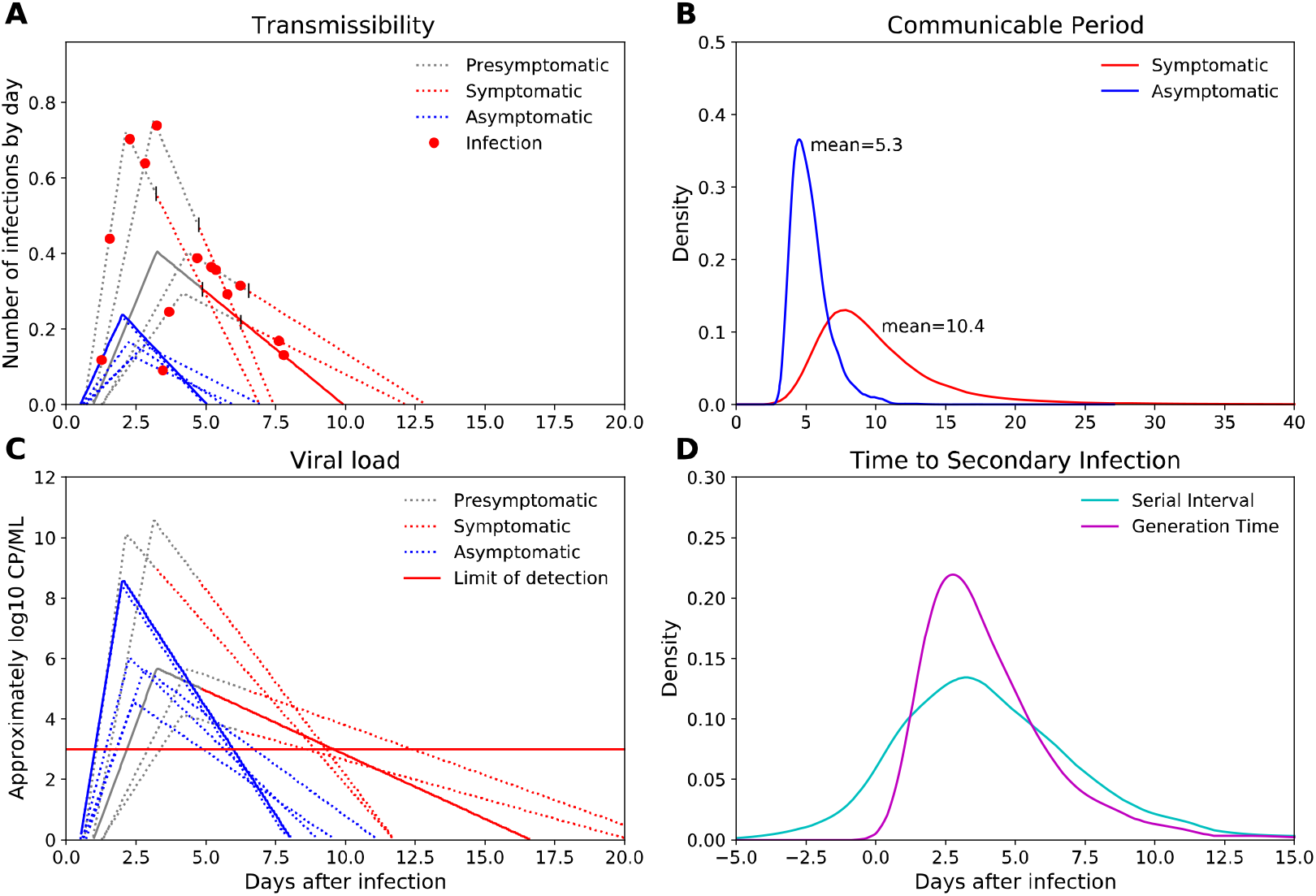
Transmissibility (A) and viral load models (B) for five symptomatic (red lines) and five asymptomatic individuals (blue lines). The x-axis represents days after infection. The y-axis represents the average number of infections per day over the course of infection. The red dots are infection events. The curves of viral load of these individuals follow the timeline of transmissibility, but with slower rate of decline, and viral load roughly following log_10_ copies/ml of virus. The horizontal line reflects the limit of detection of a test under which the sensitivity of test decreases proportionally to viral load. C) Density functions of communicable period for symptomatic and asymptomatic carriers. D). Density functions of serial interval and generation time for symptomatic carriers. All densities were estimated from 10,000 replicate simulations.

The R_e_ is the average number of secondary infections produced by an infectious person in a population. It can vary from population to population and in time based on demographics, public awareness, self-protection, and possibly viral strain (8, 9).

Estimates of COVID-19 R_e_ can be as high as 6.47 (10) and below 1 in many parts of the world. We modeled individual infectiousness with a biological component that relates to viral load and a social component that is impacted by factors such as social distancing and mask wearing. Viral load was simulated as base reproduction numbers drawn from a normal distribution with a 95% confidence interval (CI) between 1.4 and 2.8 for symptomatic cases and 0.28 to 0.56 for asymptomatic carriers (11). Social determinants of infectivity were simulated through a distancing factor that, for example, doubles the base reproduction number if a simulated carrier will interact with others intensely without social distancing after quarantine. The overall reproduction number for the population depends on the proportion of individuals who are asymptomatic carriers. The proportion of individuals who are asymptomatic carriers was drawn from a normal distribution with 95% CI of 0.1 and 0.4 (12), so on average, 25% of simulated individuals did not show any symptoms. As discussed below, the choice of reproduction numbers had no impact when determining optimal quarantine strategies.

Infected individuals underwent an incubation period that followed a lognormal distribution with a mean of 5.5 days (13). Most pre-symptomatic transmission exposure occurs 1–3 days before a person develops symptoms, and viral loads are already at their peak or declining at the onset of symptoms (14–16). Therefore, we allowed transmission probabilities to start 1 to 2 days after infection, peak before the onset of symptoms, and decline afterward. We set the period of infectivity after the onset of symptoms to follow a shifted lognormal distribution so 88% and 95% of individuals are no longer infectious after 10 and 15 days, respectively (15). The transmissibility curves for asymptomatic carriers were similar, although the overall communicable periods (periods in which carriers stay infectious) were shorter than for symptomatic cases (17, 18).

### Model of viral load

Transmission of the SARS-CoV-2 virus is caused by viral shedding from infected individuals, and the probability of infecting other individuals is related to the viral load of the infector. Viral shedding begins 5 to 6 days before the appearance of symptoms and decreases monotonically after symptom onset (19). The mean viral shedding duration is quite long, approximately 18 days for upper respiratory tract (URT) and 15 days for lower respiratory tract, with a maximum duration of RNA shedding of 83 and 59 days, respectively. However, the probability of detecting infectious the virus drops below 5% after 15.2 days post-onset of symptoms (15).

While symptomatic carriers have much higher transmissibility probabilities compared with asymptomatic carriers, recent studies found little to no difference in viral load between pre-symptomatic, asymptomatic, and symptomatic persons (19–23). However, asymptomatic carriers could have faster clearance (17, 24). For example, Yang et al (18) reported a median duration of viral shedding of 8 days (interquartile range [IQR]: 2–12) for asymptomatic and 19 days (IQR: 16–24) for symptomatic carriers. We assumed that the lower overall infectivity of asymptomatic carriers compared with symptomatic cases with similar initial viral loads was due to different immune responses to the infection. One hypothesis is that asymptomatic carriers have reduced viral transmission due to the lack of viral expectoration via cough and sneezing (25). The immune system of asymptomatic carriers could potentially fight the virus more efficiently and clear it faster.

Similar to a study by Larremore et al. (26), we modeled individual viral load patterns as piecewise functions that follow individual transmissibility curves. We extended the tails of the distributions to reflect a longer viral shedding period than communicable period and adjusted the distribution intensities, so that viral loads are proportional to the biological-driven portion of the transmissibility probabilities, and symptomatic carriers and asymptomatic carriers had similar initial viral loads (Figure 2B).

### Sensitivity of tests

We model the sensitivity of tests as both the LOD and clinical sensitivity. LOD is affected by the number of virions obtained from infected individuals based on their stage of infection. Clinical sensitivity can be affected by clinically relevant real-life situations (e.g., variations among specimen sources, the timing of sampling, and the experience of medical staff). We modeled LOD as a cutoff value where the test’s sensitivity decreases proportionally to viral load when viral load is lower than the cutoff value but stays constant when the viral load is above the cutoff value (Figure 2B).

## Results

### Model validation

We implemented and tested multiple strategies to model SARS-CoV-2 transmissibility, including using normal distributions, for both symptomatic and asymptomatic carriers.(11) We validated our models extensively using simulations that summarized the final outputs of the models, including generation time; serial intervals; and proportions of asymptomatic, pre-symptomatic, and symptomatic infections. We generated secondary infection events using a transmissibility density function, which is affected by individual reproduction number, incubation period (13), and communicable period. The average number of infection events agreed well with specified reproduction numbers. The average communicable period was 10.30 days for symptomatic cases and 5.35 for asymptomatic carriers (Figure 2C).(17, 27). Due to the long tails of the distributions used, some infected individuals had extra-long communicable periods lasting more than 20 days, which corresponds with observed outlier SARS-CoV-2 cases (27).

The distribution of generation time, defined as the time between the infection in the source and infection in the recipient for source-recipient transmission pairs, was 4.25 ± 2.75 days (mean ± standard deviation) days. This result is more spread-out, and therefore more conservative, than reported in Ferretti et al. (5.0 ± 1.9 days; Figure 2D). (11) The distribution of serial intervals, defined as days between the primary case developing symptoms and secondary case developing symptoms, from 10,000 infection events followed a normal distribution with a mean of 4.25 and standard deviation of 3.62. This result agrees with estimated mean serial intervals of 4.6 (28) and 3.96 days (29). The serial intervals followed a normal distribution with approximately 12% of pairs showing negative serial intervals, i.e., the infected individuals showing symptoms before the infector (Figure 2D). This is due to the wide spread of pre-symptomatic transmission times and is consistent with observed data (28, 30). The proportions of asymptomatic, pre-symptomatic, and symptomatic transmissions were 6%, 48%, and 44%, respectively, with the default parameters we used (12).

### Impact of testing frequency

We assumed 1) use of an RT-PCR test with a clinical sensitivity of 0.95 and 1-day turnaround time and 2) a worst-case scenario for which everyone studied was infected just prior to quarantine. We quarantined people from 1 to 14 days and tested individuals at the end of the quarantine period. Note, we defined the day of testing as the day on which test results were obtained. In practice, an RT-PCR test conducted on “day 11” would be administered at the beginning of day 10 and the result received at the beginning of day 11; whereas a rapid antigen test conducted on “day 11” would be administered at the beginning of day 11 with results available shortly thereafter.

Our simulations found testing significantly reduced the PQTR in the subsequent work-cohorts of quarantined individuals (Figure 3A). The PQTR for 14-day quarantine was reduced from 0.12% to 0.006% when people were tested before releasing them from quarantine. Using a 95% sensitivity RT-PCR test, a 9-day quarantine with a pre-release test resulted in a PQTR of 0.09%. This risk is slightly lower than what is expected with a test-free 14-day quarantine. A longer quarantine would be needed if using a less sensitive test. For example, a test with 80% sensitivity would require an 11-day quarantine to achieve similar risk mitigation. Notably, testing at the beginning of the quarantine, such as for a 1-day quarantine, had no effect because we assumed all individuals were infected recently and had no detectable viral load at the beginning of the simulation.

**Figure 3:**
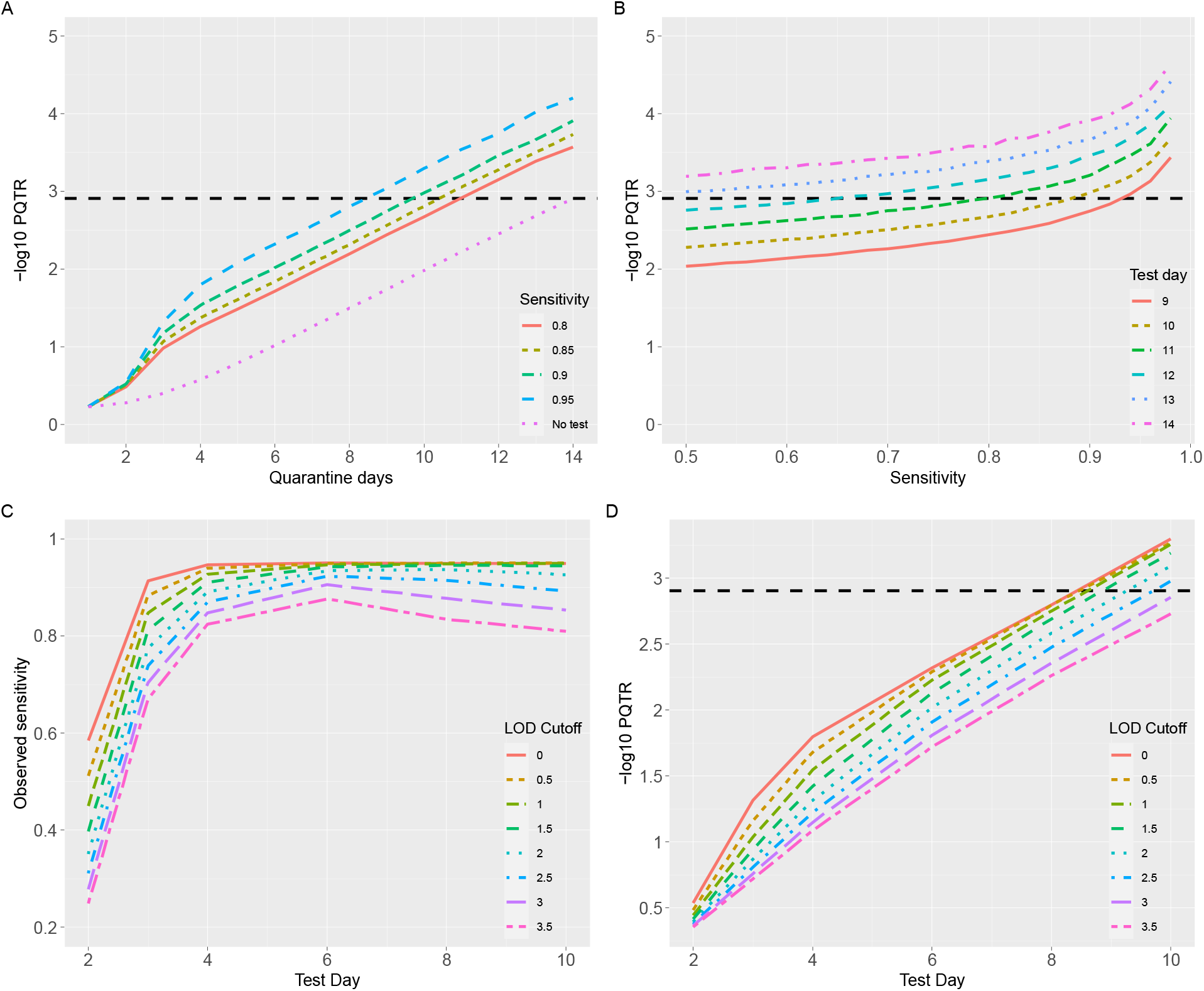
A) PQTR for quarantine from 1 to 14 days with and without testing on the day before release. B): Impact of test sensitivity on PQTR. The results are based on a 10-day quarantine with testing on the day before release. C) The observed sensitivity, derived from the proportion of false-negatives caused by the low viral load of carriers, of tests with a baseline sensitivity of 95% when performed at days 2, 3, 4, 6, 8, and 10 of quarantine. D). PQTR of the quarantine strategies with the same tests as conducted in (C) also performed at the end of quarantine with a 1-day turnaround time.

Increasing the number of tests led to shorter quarantines outperforming a test-free 14-day quarantine, mostly by reducing false-negatives through repeated testing. Assuming a 95% sensitivity RT-PCR test and 1-day turnaround time, the “quickest” way to release people from quarantine was a 6-day quarantine with tests on days 4, 5, and 6. Individuals would be released at the end of day six if all three tests were negative. The next best option was a 7-day quarantine with tests on days 4, 5 or 6, and 7 (Table 2). Adding more quarantine days or more tests would improve the performance of these strategies but would incur unnecessary costs and burden to those under quarantine.

**Table 2:** Parameters used by six simulated SARS-CoV-2 tests.

| Test Name | Peak Sensitivity | Limit of Detection | Turnaround Time | Average sensitivity | Average sensitivity on optimal test days |
| --- | --- | --- | --- | --- | --- |
| PCR95 | 95% | 1 | 1 day | 91% | 94% |
| PCR90 | 95% | 2 | 1 day | 86% | 92% |
| PCR80 | 85% | 2 | 1 day | 77% | 83% |
| PCR70 | 80% | 3 | 1 day | 66% | 75% |
| Ag80 | 95% | 3 | 1 hour | 79% | 86% |
| Ag65 | 80% | 4 | 1 hour | 59% | 66% |

Parameters used by 6 simulated SARS-CoV-2 tests. Peak sensitivity refers to sensitivity caused by factors not related to test themselves. Limit of detection refers to cutoff values for viral load below which tests have lower sensitivity proportional to viral load. Turnaround time refers to time after which tests results can be obtained. Average sensitivity is the averaged observed sensitivities if tests are performed during the communicable periods (with positive viral loads) for 10,000 random simulated
individuals with default parameters who were infected at time 0. Average sensitivity on optimal test days is the average observed sensitivity for quarantine strategies that test individuals at selected days that result in lowest PQTR.

### Impact of test sensitivity

Tests that were less sensitive increased the chance of false negatives, which resulted in higher PQTR (Figure 3A and 3B). Based on simulations of a 10-day quarantine with an RT-PCR test performed one day before release, a test with 90% sensitivity would perform comparably to a 9-day quarantine with a test with 95% sensitivity. RT-PCR has a low LOD and can detect the RNA of SARS-CoV-2 from samples with 10^3^ copies/ml (31) to as little as 10^2^ copies/ml (32). Antigen tests have a higher LOD of 10^5^–10^6^ copies/ml (33) and would have a lower chance of detecting COVID-19 carriers at the beginning and end of infection. Based on the false-negative rate, the LOD of tests had a substantial impact on clinical sensitivity at the beginning of infection, which reduced around days 5 and 6 when carriers have the highest viral load and increased at a later stage of infection. (Figure 3C).

### Impact of the proportion of asymptomatic carriers

Our quarantine strategy is more effective for asymptomatic than symptomatic carriers because asymptomatic carriers have a shorter communicable period and are less likely to infect others if not detected and released from quarantine. Therefore, the quarantine would be more effective if there are a higher percentage of asymptomatic carriers in the population. The PQTR of all quarantine strategies were higher in populations with a higher proportion of symptomatic cases. However, the relative performances of quarantine strategies were not affected by the proportion of asymptomatic cases.

### Impact of physical distancing

Reproduction numbers measure the ability of the disease to infect others. The effective transmissibility, R_e,_ indicates the actual transmissibility accounting for behaviors, such as mask use or social distancing. In our model, real-time R_e_ is zero during quarantine and positive based on working environments after release. We used distancing factors to model social distancing in the working environments, which affect the probabilities that carriers infect others, but not their viral loads and probabilities to be detected.

As shown in Supplementary Table 1, the PQTR of both test-free and test-assisted quarantine strategies were higher if released individuals work in an enclosed environment without social distancing than in a more protected environment with social distancing and mask use. However, compared to a test-free quarantine, in which all asymptomatic carriers are released and become more infectious in environments with less distancing, test-assisted quarantine strategies detect most carriers and release them only after they are no longer infectious. The PQTR of test-assisted quarantines increases slower than test-free quarantines when simulating scenarios with progressively decreasing social distancing practices. Most of the test-assisted quarantine strategies (with shorter quarantine intervals) outperformed the test-free quarantines (Supplementary Table 1). Therefore, test-assisted strategies are more effective than test-free quarantines for the assembling of individuals without social distancing.

### Impact of individual viral loads

In contrast to physical distancing that affects only post-quarantine transmission probabilities, individual viral loads impact both the probability that a carrier is detected during quarantine, and their transmission probability after the quarantine period. Using a strategy that tests individuals 24-hours before the end of a 10-day quarantine, we simulated five groups of individuals with varying overall viral loads (from ½ to 2 times) and estimated observed test sensitivities and PQTR for four tests with varying sensitivity and limits of detection. Because test sensitivities plateau when viral loads exceed the limit of detection of tests, the impact of viral loads on post-quarantine transmission probabilities exceeds its impact on test sensitivities, leading to roughly linear increase of PQTR with increased viral loads. This trend is appreciable in Figure 4.

**Figure 4:**
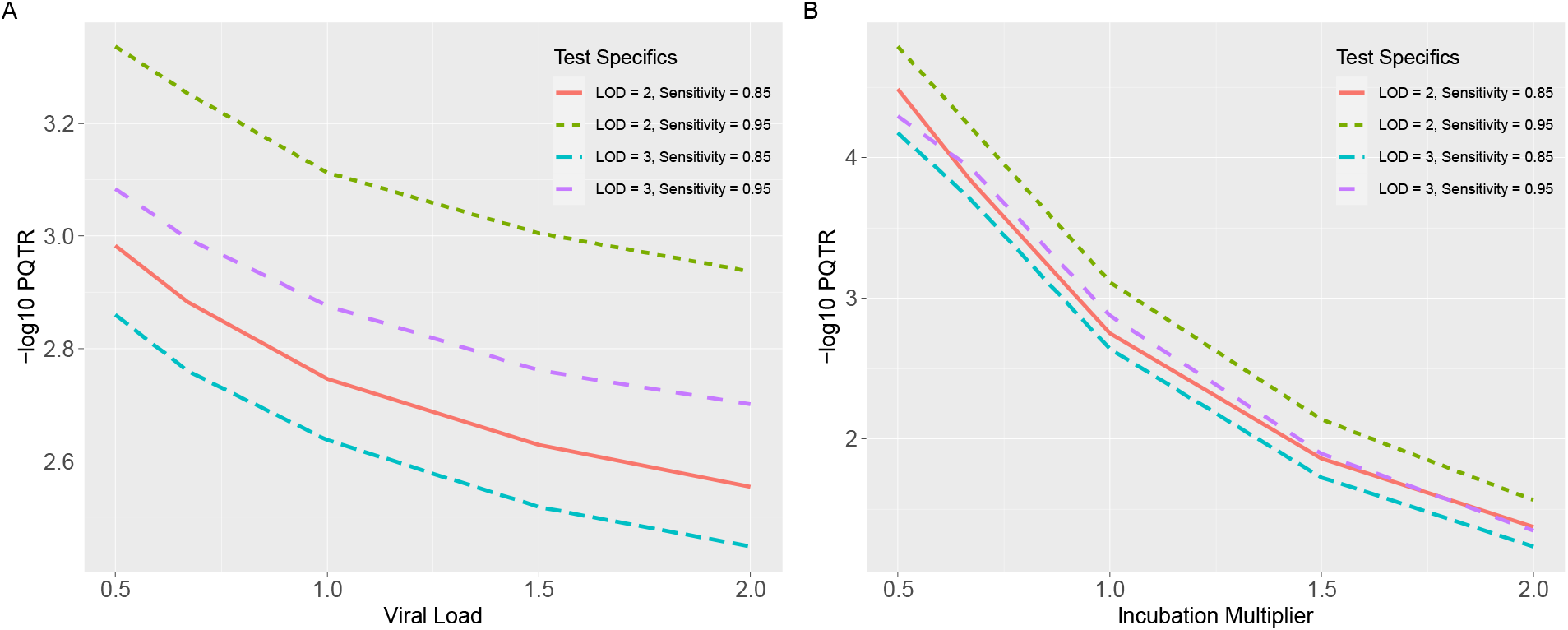
A) PQTR for a 10-day quarantine strategy with four different tests administered on the day before release, for people with viral load that are 1/2, 2/3, 1, 1.5, and 2.0 times of the average viral load of the simulated population. B) PQTR for the same quarantine strategies and tests, for people with 1/2, 2/3, 1, 1.5, and 2.0 times of the average incubation period of the simulated population.

### Impact of incubation periods

The length of the SARS-CoV-2 incubation period, or the time from exposure to the development of symptoms, determines the likelihood that the carrier shows symptoms during a quarantine intervention. Incubation period length further influences the viral load and communicable period of a carrier, and therefore their post-quarantine transmission probabilities. In Figure 4 we present our results of varying the relative incubation length of viral infection from ½ to 2 times in duration. It can be appreciated that PQTR increases exponentially, independent of testing strategy, as the incubation period lengthens.

### Optimal strategies for widely used SARS-CoV-2 tests

PCR-based tests can detect the existence of SARS-CoV-2 RNA from upper respiratory tract samples of recovered patients for up to 12 weeks, even when the replication-competent virus was not detected more than three weeks after symptom onset (34, 35). The clinical sensitivity of SARS-CoV-2 tests is affected by test design, the viral loads of infected individuals, and many human and non-human factors. Therefore, although sensitivity for COVID-19 tests that are derived from testing samples with the SARS-CoV-2 virus in a lab environment is generally very high (usually >95%), the real-world performance of these tests can be significantly poorer (36). Reported clinical sensitivities include 71% (37), 86% (38), 82% in a more recent retrospective study (39), and a systematic review of 34 studies involving 12,057 patients showed test sensitivities ranging from 42% to 98% with a median of 89% (40). In addition to cross-study differences, another meta-analysis of 1,330 samples from 7 previously published studies showed median sensitivities that start from 0% at the onset of infection, 34% at day 4, peak at 80% one day after the onset of symptoms, and decrease to 34% at day 21 (41). More recent studies, however, attributed the lower sensitivity of the earlier studies partly to the use of oropharyngeal instead of nasopharyngeal swabs and found that RT-PCR tests generally have more than 90% sensitivity (42).

Test-assisted quarantine strategies offset the increased PQTR caused by shorter quarantine days through the identification of virus carriers before they develop symptoms. Effective test-assisted quarantine strategies test individuals at the peak of their viral loads at which tests are most sensitive and use repeated testing to further reduce false negatives and increase sensitivity. To evaluate the performance of commonly used SARS-CoV-2 tests, we modeled six SARS-CoV-2 tests that include a) a best-case scenario in which high-sensitivity RT-PCR tests are applied to samples collected from nasopharyngeal or mid-turbinate swabs, b) a common scenario with a regular RT-PCR test on anterior nasal swabs that misses positives due to lower viral loads (Ct >30–32), c) two RT-PCR tests with lower sensitivities due to poor quality of tests, protocol, or administration of tests, and d) two antigen tests with lower sensitivity, but faster turnaround time (1 hour) and lower cost (Table 2). Due to variations of viral loads at different stages of infection (41), we model the sensitivities of the tests with a baseline peak sensitivity value that reflects common factors unrelated to the test themselves and LOD cutoff values that reflect analytic sensitivities of the tests. The observed sensitivity of these tests varies over the course of the infection (Figure 3C). The average sensitivities, if the tests are applied every day during the communicable periods with positive viral loads, range from 66% to 91% for four RT-PCR tests and 59% to 79% for two antigen tests.

Assuming default parameters for R_0,_ no social distancing after quarantine, and proportion of asymptomatic carriers in the population, we evaluated the performance of quarantine strategies with all possible combinations of testing schedules using these six tests for those undergoing quarantine with either simultaneous or mixed onset of infections (Table 3, 4, and Supplementary Table 1). We found numerous quarantine strategies that performed better than a test-free 14-day quarantine and reported only those with the shortest quarantine or smallest number of tests. Our results indicate that quarantine on people with mixed onset of infection has much lower PQTR (generally 1/10) and a wider choice of safe test-assisted quarantine strategies compared with quarantine with the simultaneous onset of infection (Supplementary Table 1).

**Table 3:**
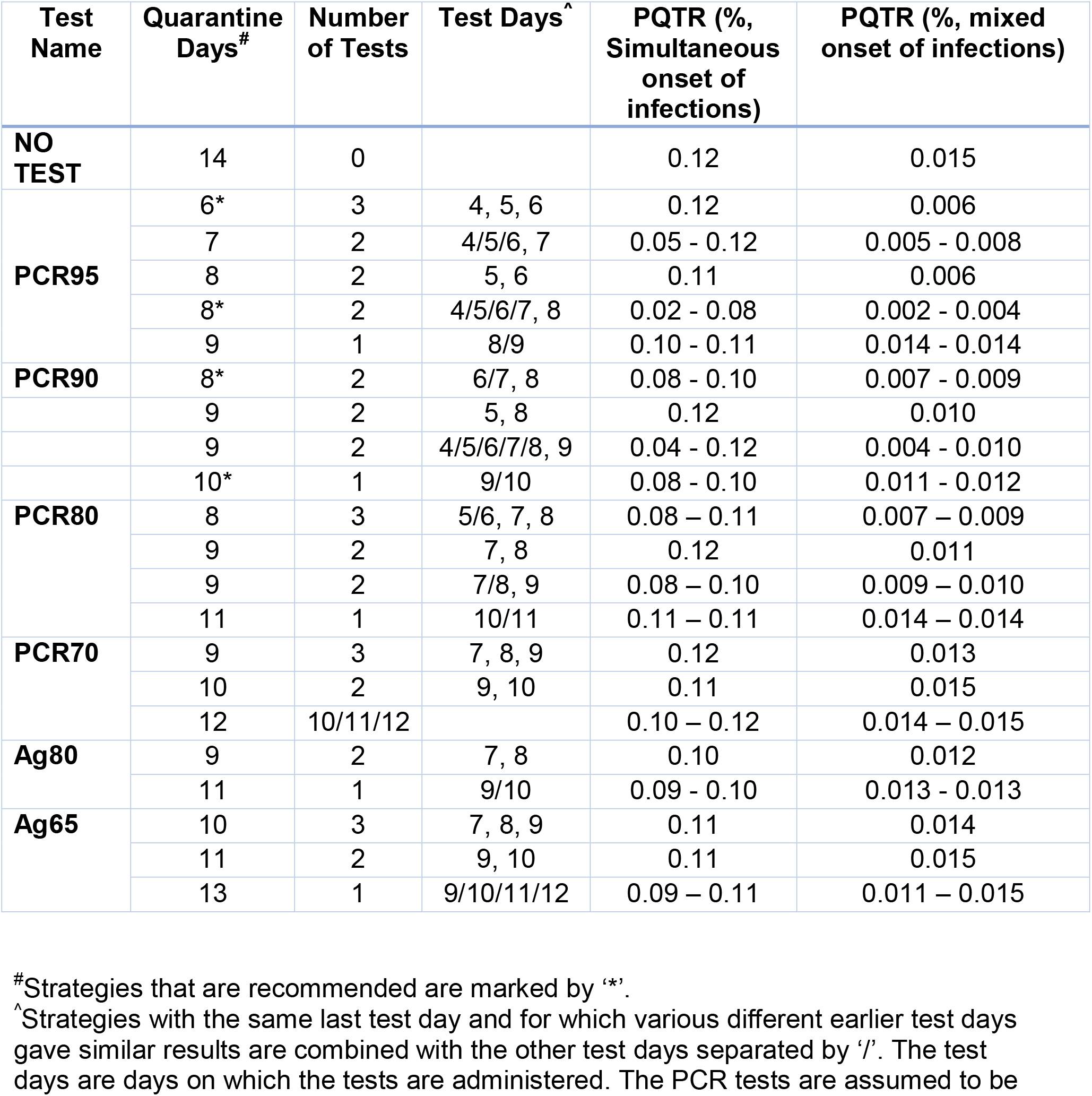
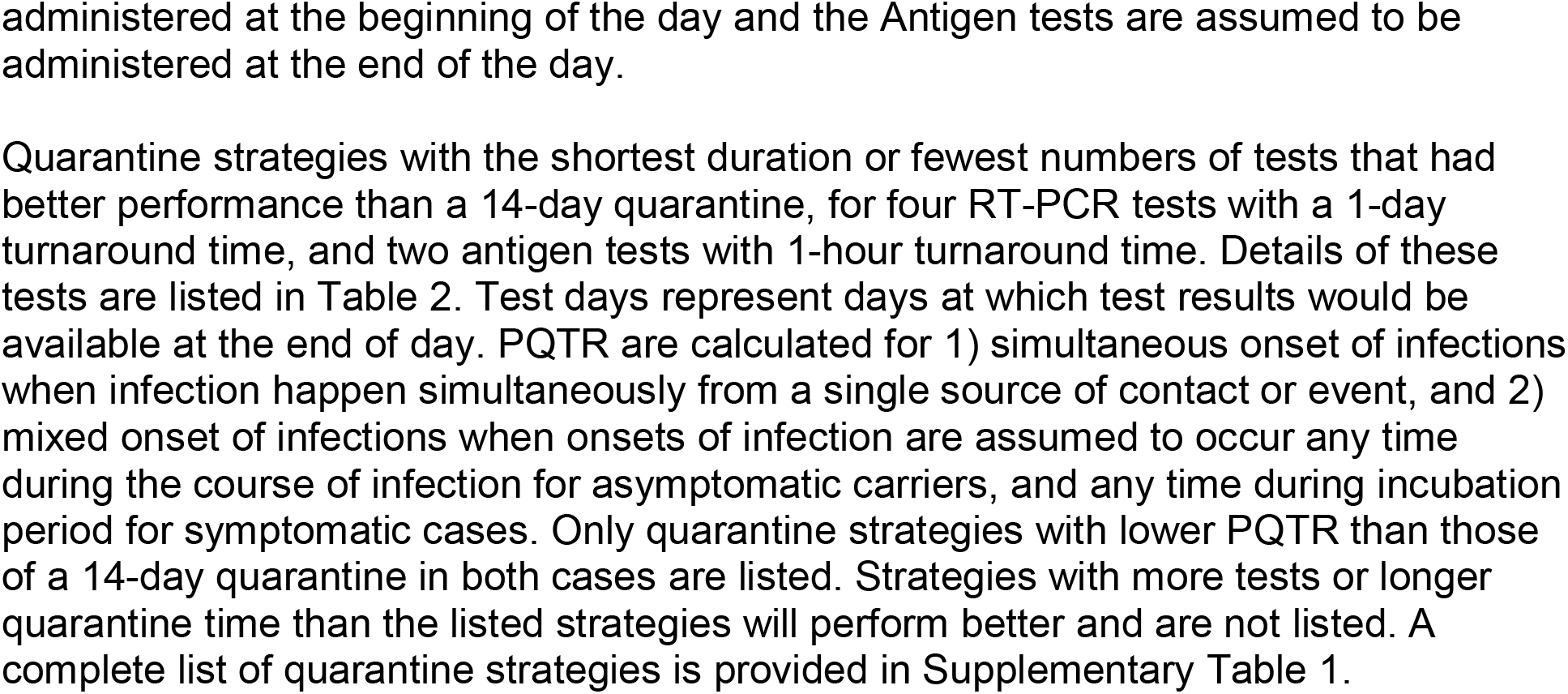
Optimal quarantine strategies with the shortest duration or fewest numbers of tests that had better performance than a 14-day quarantine.

| Test Name | Quarantine Days<sup>#</sup> | Number of Tests | Test Days<sup>^</sup> | PQTR (%<br>Simultaneous<br>onset of<br>infections) | PQTR (%<br>mixed<br>onset of infections) |
| --- | --- | --- | --- | --- | --- |
| **NO TEST** | 14 | 0 |  | 0.12 | 0.015 |
| **PCR95** | 6* | 3 | 4, 5, 6 | 0.12 | 0.006 |
|  | 7 | 2 | 4/5/6, 7 | 0.05 - 0.12 | 0.005 - 0.008 |
|  | 8 | 2 | 5, 6 | 0.11 | 0.006 |
|  | 8* | 2 | 4/5/6/7, 8 | 0.02 - 0.08 | 0.002 - 0.004 |
|  | 9 | 1 | 8/9 | 0.10 - 0.11 | 0.014 - 0.014 |
| **PCR90** | 8* | 2 | 6/7, 8 | 0.08 - 0.10 | 0.007 - 0.009 |
|  | 9 | 2 | 5, 8 | 0.12 | 0.010 |
|  | 9 | 2 | 4/5/6/7/8, 9 | 0.04 - 0.12 | 0.004 - 0.010 |
|  | 10* | 1 | 9/10 | 0.08 - 0.10 | 0.011 - 0.012 |
| **PCR80** | 8 | 3 | 5/6, 7, 8 | 0.08 – 0.11 | 0.007 – 0.009 |
|  | 9 | 2 | 7, 8 | 0.12 | 0.011 |
|  | 9 | 2 | 7/8, 9 | 0.08 – 0.10 | 0.009 – 0.010 |
|  | 11 | 1 | 10/11 | 0.11 – 0.11 | 0.014 – 0.014 |
| **PCR70** | 9 | 3 | 7, 8, 9 | 0.12 | 0.013 |
|  | 10 | 2 | 9, 10 | 0.11 | 0.015 |
|  | 12 | 10/11/12 |  | 0.10 – 0.12 | 0.014 – 0.015 |
| **Ag80** | 9 | 2 | 7, 8 | 0.10 | 0.012 |
|  | 11 | 1 | 9/10 | 0.09 - 0.10 | 0.013 - 0.013 |
| **Ag65** | 10 | 3 | 7, 8, 9 | 0.11 | 0.014 |
|  | 11 | 2 | 9, 10 | 0.11 | 0.015 |
|  | 13 | 1 | 9/10/11/12 | 0.09 – 0.11 | 0.011 – 0.015 |

<sup>#</sup>Strategies that are recommended are marked by ‘\*’.
<sup>^</sup>Strategies with the same last test day and for which various different earlier test days gave similar results are combined with the other test days separated by ‘/’. The test days are days on which the tests are administered. The PCR tests are assumed to be
administered at the beginning of the day and the Antigen tests are assumed to be administered at the end of the day.

**Table 4:** Optimal quarantine strategies with the shortest duration or fewest numbers of tests that had better performance than a 10-day quarantine.

| Test Name | Quarantine Days<sup>#</sup> | Number of Tests | Test Days<sup>^</sup> | PQTR (%<br>Simultaneous Onset of Infections) | PQTR (%<br>Mixed Onset of Infections) |
| --- | --- | --- | --- | --- | --- |
| **NO TEST** | 10 | 0 |  | 1.01 | 0.154 |
| **PCR95** | 5* | 2 | 3/4, 5 | 0.47 - 0.75 | 0.028 - 0.047 |
|  | 6 | 1 | 5/6 | 0.60 - 0.87 | 0.097 - 0.110 |
| **PCR90** | 6* | 1 | 6 | 0.96 | 0.110 |
| **PCR80** | 6 | 2 | 4/5, 6 | 0.80 - 0.84 | 0.078 – 0.107 |
|  | 8 | 1 | 6/7/8 | 0.57 – 0.83 | 0.085 – 0.105 |
| **PCR70** | 7 | 2 | 5/6, 7 | 0.69 – 0.89 | 0.091 – 0.108 |
|  | 8 | 1 | 8 | 0.74 | 0.134 |
| **Ag80** | 6* | 2 | 4, 5 | 0.94 | 0.089 |
|  | 7* | 1 | 6 | 0.81 | 0.117 |
| **Ag65** | 7 | 3 | 3/4, 5, 6 | 0.80 – 0.93 | 0.094 – 0.109 |
|  | 8 | 2 | 4/5, 6 | 0.80 – 0.96 | 0.103 – 0.123 |
|  | 8 | 2 | 4/5/6, 7 | 0.67 – 0.71 | 0.077 – 0.128 |
|  | 9 | 1 | 6/7/8 | 0.63 – 0.87 | 0.097 – 0.120 |

Quarantine strategies with the shortest duration or fewest numbers of tests that had better performance than a 10-day quarantine, for four RT-PCR tests with a 1-day
turnaround time and two Antigen tests with a 1-hour turnaround time. Details of these tests are listed in Table 2.

Considering only strategies that outperform a standard 14-day quarantine for individuals with both simultaneous and mixed onset of infections, a single RT-PCR test administered one or two days before the end of quarantine could reduce the duration of quarantine to 10 days. Two RT-PCR tests administered on days 6 or 7 and then day 8 could reduce the duration to 8 days. In cases where the shortest quarantine is needed, a 6-day quarantine with tests on days 4, 5, and 6 using a highly sensitive RT-PCR test can be justified. Longer quarantine times are required for tests with lower sensitivity. For example, two more days of quarantine would be needed if the test PCR70 is used (Table 2, 3).

Due to their high LOD, antigen tests are less sensitive in detecting the presence of the SARS-CoV-2 virus at early and later stages of infections. A single antigen test with test administered on day 9 or 10 could reduce the quarantine to 11 days. Persons in quarantine who tested negative at the beginning of days 8 and 9 could be released at the beginning of day 10 if they do not show any symptoms (a 9-day quarantine with tests on days 7 and 8). Longer quarantines would be needed for antigen tests with lower sensitivity. Shorter quarantines, even with daily antigen testing, did not achieve the same level of safety as a test-free 14-day quarantine.

For applications in which higher residual risks could be tolerated, or be mitigated by, for example, strict physical distancing and post-quarantine symptom monitoring, a 10-day test-free quarantine with a PQTR of 1.05% is recommended by the December 2020 guideline from the US CDC. A number of test-assisted quarantines, including 6-day quarantine with one RT-PCR test and 5-day quarantine with two RT-PCR tests, provide lower PQTR than 10-day test-free quarantine, which was suggested by the U.S. CDC. Quarantine strategies with testing at the beginning and end of quarantine incur residual risk. As a real-world example, the New York travel recommendation resembles an 8-day quarantine strategy with testing on days 1 and 8, The PQTR of this strategy is 0.024%, which is 60% higher than a 14-day quarantine even if we assume that traveling adds no additional risk of infection. We report in Table 4 those strategies with either the shortest quarantine time or smallest number of tests.

## Discussion

Using our flexible and publicly available COVID-19 Python-coded simulator (https://ictr.github.io/covid19-outbreak-simulator/), we modeled the efficacy of foreshortened quarantine strategies with and without the concurrent implementation of Covid-19 testing. We evaluated the efficacy of these test-assisted quarantines through modulating the simulated test sensitivities and turnaround times of carriers at different stages of infection. Further, we modeled the effect of specific implementation scenarios, based on our existing knowledge on the progression of SARS-CoV-2 infection and performance of common RT-PCR and antigen tests. We hypothesized that COVID-19 testing would allow for a shorter quarantine than a test-free 14 or 10-day quarantine strategy. The simulations identified specific shortened quarantined strategies that could be effective.

Using a relatively sensitive RT-PCR test and current lab protocols, ending a quarantine at day 10 with a single RT-PCR test performed one or two days before release outperformed a test-free 14-day quarantine for both simultaneous and mixed-onset COVID-19 exposures (Table 3). Exceptionally short quarantines, including a 6-day quarantine, were justifiable if repeated high sensitivity RT-PCR testing was performed on days 4, 5, and 6, and even shorter quarantine strategies could be used for applications with higher tolerance to PQTR (Table 4). Quarantining individuals with mixed exposure instances yielded a wider range of effective quarantine strategies than did a cohort of simultaneously exposed individuals. Epidemiologically, we found the average communicable period for persons with symptomatic cases of COVID-19 was 10.30 days compared with 5.35 for asymptomatic carriers.

We estimated PQTR of test-assisted quarantined based on a mixed population with varying properties such as viral load, incubation period, communicable period, proportion of asymptomatic carriers. However, age (43) (44), male gender (45), elevated risk profile (46) are all associated with the poor prognosis of COVID-19 diseases (47) and mortality (48) so different quarantine strategies might be needed for cohorts consisting with seniors, males, or people with multiple complications.

The false-positive rates of RT-qPCR tests are associated with patient age, sex, and time since diagnosis, although the association is best explained by individual viral loads (49). Since the average differences in viral loads and resulting differences in false-negative rates and PQTR between male and females are relatively small (∼ 5% in false negative rates) compared to differences between people under and over the age of 40 (∼20% in false negative rates) (49), a more stringent test-assisted quarantine strategy should be used for cohorts with predominantly people over the age of 40.

Individuals with multiple co-morbidities tend to have shorter incubation periods, higher viral loads, and worse prognoses. We found each of these factors increases the efficiency of test-assisted quarantines (as shown in Figure 4); therefore, we suggest that our recommended quarantine strategies should be applicable to cohorts with elevated risk profiles.

The emergence and wide spread of <u>new variants</u> of the SARS-CoV-2 virus pose new challenges to effective application of quarantine strategies. For example, the 614G variants propagate more quickly than the wide-type (50) and preliminary studies suggest that the 501Y.V2 variant is associated with a higher viral load. The variants might also evade detection by specific viral diagnostic tests although most commercial RT-PCR tests have multiple targets to detect the virus and should continue to work. Although there is insufficient data for us to develop variant-specific quarantine strategies, we recommend the use of more stringent quarantine strategies in areas with high prevalence of these new variants.

Strengths of our approach include repeated and systematic simulation comparisons that allow us to confidently draw conclusions regarding the efficacy of these quarantine models based on our accurate probabilistic modeling of person-to-person COVID-19 spread in well-mixed populations. Strengths of our model include effective implementation and accurate modeling of COVID-19 spread in diverse real-world scenarios (7). A potential limitation of our modeling technique is our use of probabilistic modeling. We simulated the occurrence of infection and recovery events at random based off measured parameter probabilities of COVID-19 spread within a network or between networks. This limits the contribution of geographical constraints or non-random social interaction. Further, we found lengthened incubation periods diminishes the efficacy of quarantine strategies. Therefore, it is critically important to understand variations in incubation period, especially for test-assisted quarantine strategies with shorter quarantine durations. Since individuals with shorter incubation periods are more likely to show symptoms during quarantine (Figure 4b), individuals at the tail of the distribution of incubation period contribute to the majority of cases who show symptoms and infect others after quarantine, and to the PQTR of our presented quarantine strategies.

The literature on foreshortened and test-assisted quarantine strategies is sparse. Only two investigations similar to ours have been conducted, and currently, neither have been peer reviewed. The first study extrapolated epidemiological parameters of COVID-19 from a real-world dataset from COVID-19 testing of offshore oil rig employees (51). The second used a model to capture transmission associated with air travel and develop quality strategies to mitigate the entry of COVID-19 into new regions (52). Both studies had limitations similar to ours. They had to estimate epidemiological COVID-19 parameters from limited real-world datasets, and they had to create probabilistic models to test the efficacy of COVID-19 testing in their respective scenarios. Our base model has a distinct advantage due to its generalizability, unique flexibility, and easy programming with command line prompts to simulate a wide range of scenarios. We have also iteratively tested the efficacy of the quarantine strategies in a broad range of scenarios. Collectively, these model features ensure our simulations are robust.

A pressing need exists for evidence-based COVID-19 mitigation strategies. Currently, deviations from a 14-day post-exposure quarantine have little substantiating evidence supporting their utility. Despite a lack of systematic simulation modeling, shortened and test-assisted quarantine strategies are being implemented out of logistical necessity. Our work fills a clear gap in our understanding of quarantine strategy efficacy by comprehensively simulating alternative isolation strategies. The increased efficacy we identified using test-assisted quarantine strategies should also be appreciated beyond their ability to shorten the overall quarantine. A major difference between test-free and test-assisted quarantine is that test-assisted quarantines identify asymptomatic carriers and carriers who have recovered but are still shedding, i.e., those who would pass test-free quarantine unnoticed. Therefore, the 10-day test-assisted quarantine we identified or the even shorter quarantines with repeated RT PCR testing demonstrate superior efficacy at mitigating viral spread then a test-free quarantine process. This underscores the value of test-assisted quarantine as it reduces the probability of subsequent quarantine failure while also releasing uninfected individuals from quarantine earlier.

We comment that this recommendation is in line with the recent recommendations from the CDC. For example, the CDC recommended 10-day and 7-day quarantines with a RT-PCR test performed within 48 hours before the end of the quarantine, which are similar to our recommended 9-day and 6-day quarantines with tests performed within 24 hours before the end of the quarantine. Due to the wide range of test sensitivities, the CDC reported wide ranges of PQTR and recommended, among other methods, continued monitoring of symptoms after releasing from quarantine to mitigate the risk caused by the use of tests with lower sensitivities.

Test-assisted quarantines require the administration of SARS-CoV-2 tests. The costs of testing depend on the size of cohort, number, and type of tests, and could outweigh the benefits of reduced quarantine time. We recommend optimal test-assisted quarantine strategies for different types of tests with one or more tests during quarantine and allow the use of most cost-effective strategies for particular applications. Notably, testing cohorts with uninfected individuals, especially repeated testing of large cohorts, generates false positives. RT-PCR tests have very high specificity (between 99.9% and 99.95%) and produce false-positives only through human errors during pre-analytical and analytical processes. These values are higher than those of lower specificity antigen tests (e.g., <u>98.5% for BinaxNow</u>). Therefore, <u>the F.D.A. recommends</u> positive tests obtained by antigen tests are confirmed with an RT-PCR test. Unless a physician suggests otherwise, people who test positive should complete a 10-day isolation as recommended by the CDC

Effective vaccines that can generate host immunity to the novel SARS Co V2 virus. A clear direction for future research involves modeling viral spread in populations with varying degrees of vaccine-induced immunity. Specifically, questions that remain unaddressed include the efficacy of the quarantine strategies we present given known vaccination status, the timing of recent vaccination, and the vaccination status of the population or subpopulations of interest.

## Conclusion

Our findings indicate that the current 14-day quarantine approach is excessively conservative, and the duration of quarantines could be significantly reduced if testing is performed during the quarantine period (Table 3 and 4). To achieve a PQTR of 0.1%, comparable to a 14-day test-free quarantine, one RT-PCR test or two antigen tests could reduce the quarantine time to 10 days. More tests can further reduce the length of duration, and a 6-day quarantine can be justified with three highly sensitive RT-PCR tests. Even shorter quarantine strategies could be employed if a higher PQTR of 1%, comparable to a 10-day test-free quarantine is tolerated. We modeled uninterrupted quarantine in isolation with tests with relatively short turnaround time. If in practice, the turnaround time of a test is longer than one day, the quarantine should be extended to wait for the results of the tests.

The effectiveness of test-assisted quarantine depends on the sensitivity of tests. Because SARS-CoV-2 tests have lower sensitivity when administered at an early stage of infection with low viral loads, quarantines shorter than 6 days will be ineffective even with daily SARS-CoV-2 testing. Quarantine strategies with testing at the beginning and end of quarantine or traveling with pre-travel testing could incur residual risk. Tests with lower than 85% clinical sensitivity are unlikely to reduce the length of quarantine, and we do not recommend using test-assisted quarantine with novel tests with unknown clinical sensitivities.

## Data Availability

The simulation program (covid19-outbreak-simulator), that is used for all simulations for this study is publicly available at https://github.com/ictr/covid19-outbreak-simulator under a BSD 3-clause license. All simulated scenarios and their results will be made available in the Applications section of the homepage of the simulator.

## Acknowledgements

The authors appreciate a scientific editor, Kat H. Sippel, PhD, E.L.S. for her assistance in the preparation of this manuscript. Dr. Amos is a Research Scholar of the Cancer Prevention Research Institute of Texas and supported by RR170048 and National Institutes of Health Award Number 1OT2HL158258-01. Rowland Pettit is a fellow under the Training in Precision Environmental Health Sciences (TPEHS) Program (N.I.H. Grant No. T32ES027801). The authors would like to thank BRASS: Baylor Research Advocates for Student Scientists for their support of Rowland Pettit.

## Author contributions

B.P. designed the study, implemented the simulations, and drafted the manuscript. W.Z. executed simulations and analyzed results. B.P., W.Z., and P.Y. verified the data and results. P.Y., PM, AG, and J.M. provided real-world data, industrial practices, and government regulations for COVID-19 quarantines. C.A. supervised the study. P.Y., R.P., and C.A. contributed substantially to the manuscript. All authors reviewed and agreed with the content of the manuscript.

## Data sharing statement

The simulation program (covid19-outbreak-simulator) used for all simulations in this study is publicly available at https://github.com/ictr/covid19-outbreak-simulator under a BSD 3-clause license. All simulated scenarios and their results will be made available in the “Applications” section of the homepage of the simulator.

## Supplementary Tables

**Supplementary Table 1:** PQTR of more than 10,000 simulated quarantine strategies for 4 RT-PCR tests and 2 Antigen tests listed in Table 2. PQTR are calculated for two scenarios when 1) onset of infection happens simultaneously from a single source of contact or event, or 2) onset of infection is unknown and assumed to be any time during the course of infection for asymptomatic carriers and during incubation period for symptomatic cases.

## Transparency declaration

The lead author, Christopher Amos, affirms that the manuscript is an honest, accurate, and transparent account of the study being reported; that no important aspects of the study have been omitted; and that any discrepancies from the study as planned (and, if relevant, registered) have been explained. All approaches used in this study are available online at (https://ictr.github.io/covid19-outbreak-simulator/)

